# Secreted protein profiling of human aortic smooth muscle cells identifies vascular disease associations

**DOI:** 10.1101/2023.11.10.23298351

**Authors:** Rédouane Aherrahrou, Ferheen Baig, Konstantinos Theofilatos, Dillon Lue, Alicia Beele, Tiit Örd, Minna U Kaikkonen, Zouhair Aherrahrou, Qi Cheng, Saikat Ghosh, Santosh Karnewar, Vaishnavi Karnewar, Aloke Finn, Gary K. Owens, Michael Joner, Manuel Mayr, Mete Civelek

## Abstract

**Background:** Smooth muscle cells (SMCs), which make up the medial layer of arteries, are key cell types involved in cardiovascular diseases (CVD), the leading cause of mortality and morbidity worldwide. In response to microenvironment alterations, SMCs dedifferentiate from a “contractile” to a “synthetic” phenotype characterized by an increased proliferation, migration, production of extracellular matrix (ECM) components, and decreased expression of SMC-specific contractile markers. These phenotypic changes result in vascular remodeling and contribute to the pathogenesis of CVD, including coronary artery disease (CAD), stroke, hypertension, and aortic aneurysms. Here, we aim to identify the genetic variants that regulate ECM secretion in SMCs and predict the causal proteins associated with vascular disease-related loci identified in genome-wide association studies (GWAS).

**Methods:** Using human aortic SMCs from 123 multi-ancestry healthy heart transplant donors, we collected the serum-free media in which the cells were cultured for 24 hours and conducted Liquid chromatography-tandem mass spectrometry (LC-MS/MS)-based proteomic analysis of the conditioned media.

**Results:** We measured the abundance of 270 ECM and related proteins. Next, we performed protein quantitative trait locus mapping (pQTL) and identified 20 loci associated with secreted protein abundance in SMCs. We functionally annotated these loci using a colocalization approach. This approach prioritized the genetic variant rs6739323-A at the 2p22.3 locus, which is associated with lower expression of LTBP1 in SMCs and atherosclerosis-prone areas of the aorta, and increased risk for SMC calcification. We found that LTBP1 expression is abundant in SMCs, and its expression at mRNA and protein levels was reduced in unstable and advanced atherosclerotic plaque lesions.

**Conclusions:** Our results unravel the SMC proteome signature associated with vascular disorders, which may help identify potential therapeutic targets to accelerate the pathway to translation.

## INTRODUCTION

Smooth muscle cells (SMCs) play a vital role in the blood vessel wall by controlling blood pressure and blood flow distribution and sustaining vascular structural integrity. In pathophysiological conditions, SMCs dedifferentiate from a “contractile” phenotype to various phenotypes that can be detrimental or beneficial for plaque pathogenesis^1,2^. The latter includes a myofibroblast phenotype present within the protective fibrous cap that is characterized by increased proliferation, migration, production of extracellular matrix (ECM) components, and a decrease in SMC-specific contractile markers^3,4^. Examples of detrimental SMC phenotypic changes include their transition to pro-inflammatory foam cells or osteochondrogenic phenotypes likely to contribute to plaque calcification. Indeed, there is compelling evidence that SMC phenotypic transitions play an essential role in several cardiovascular diseases, including atherosclerosis, hypertension and aortic aneurysms^5^.

ECM, mainly produced by SMCs, is a key component of the fibrous cap of atherosclerotic lesions, which is critical for plaque stability. SMC-derived ECM also plays a regulatory and structural role in the vascular network^6^. It is a composite of several proteins that form structures connecting cells within the network^6^. ECM consists of many macromolecules, including collagen, glycoproteins, proteoglycans, and elastin, which confer tensile strength and viscoelasticity to the arterial wall^7^. The ECM synthesis can play beneficial or detrimental roles during pathological processes in the arteries. For example, high ECM synthesis in the atherosclerotic plaque leads to plaque stabilization in advanced stages but also contributes to luminal stenosis in early stages^8–10^. Degradation of ECM promotes the weakening of plaque by increasing its vulnerability to rupture that leads to myocardial infarction or stroke^7^. In addition, ECM dysregulation is associated with high blood pressure^6^ and aortic aneurysms^11^.

Genome-wide association studies (GWAS) have linked genetic loci with cardiovascular traits^12^. However, the molecular mechanisms of most of these loci are unknown. In addition, most of these loci map to non-coding regions of the genome, pointing to their effect on regulating gene expression. Integrating GWAS and quantitative trait locus (QTL) mapping studies can identify potential causal genes and proteins affecting complex cardiovascular traits. For example, the identification of ECM proteins whose abundance or synthesis is regulated by myocardial infarction-associated GWAS loci can provide a way to determine new therapeutic targets or biomarkers for plaque stability.

To date, there are no reliable SMC ECM biomarkers that can be used to distinguish patients at high risk for atherosclerotic plaque rupture^13^. For example, other than the histological appearance of the lesions, there are no biomarkers that can be used to distinguish stable and unstable atherosclerotic plaques. Although some inflammatory markers such as C-reactive protein are high in persons at risk for myocardial infarction, they lack the specificity required for the reliable distinction between the “vulnerable” and “stable” plaques. Therefore, new approaches are needed to identify ECM biomarkers associated with the cardiovascular disease stages or novel targets for treatment. Here, we hypothesized that a subset of the genomic loci associated with vascular diseases regulate ECM synthesis and secretion from the SMCs to affect the disease process. We used a multi-omics approach to identify genetic variants that are associated with secreted ECM proteins from aortic SMCs isolated from 123 healthy and ancestrally diverse heart transplant donors. This allowed us to identify vascular disease-associated loci that perturb SMC-secreted protein abundance.

## MATERIALS and METHODS

### Cell culture

We isolated primary SMCs of ascending aorta from 151 healthy and multi-ancestry heart transplant donors (118 males and 33 females) as described previously^14,15^. The SMC isolation procedure did not employ enzymes or other additives. Instead, we gently removed the endothelial cells (ECs) and followed it with delicate rubbing using Q tips to eliminate any residual ECs and ECM. Next, we cut the medial layer of the tissue into small squares and subsequently placed them onto plates coated with collagen. We gently positioned a sterile coverslip on top to secure the tissue in place, creating a microenvironment rich in growth factors around the explant, as observed under microscopic examination. The coverslip facilitated tissue attachment to the plate and potentially facilitated the release of growth factors from the explant. After the migration of SMCs from the tissue, we removed the coverslip and conducted rigorous washing to ensure thorough cell cleaning. We cultured the cells in Smooth Muscle Cell Basal Medium (SmBM, CC-3181, Lonza) supplemented with Smooth Muscle Medium-2 SingleQuots Kit (SmGM-2, CC-4149, Lonza) (complete media) and 5% fetal bovine serum (FBS) until 90% confluence. We then replaced the media with complete media without FBS. We collected the media after 24 hours for proteomics profiling. The Institutional Review Board of the University of Virginia approved this study.

### Genotyping, ancestry determination, and sample swap identification

We genotyped the donors using the Illumina Multi-Ethnic Global genotyping array for 1.8 million single nucleotide polymorphisms (SNPs). We pruned the SNPs based on call rate (< 2%), Hardy-Weinberg equilibrium (P_HWE_<1×10^-6^), and minor allele frequency (< 5%), and imputed non-genotyped SNPs using the 1000 Genomes Phase 3 reference panel^16^. After removing the SNPs with minor allele frequency less than 5%, imputation quality (MaCH r^2^ > 0.3) and significant deviation from Hardy-Weinberg equilibrium (P-value ≤ 10^-6^) we were left with ∼6.1 million SNPs. We clustered the donor genotypes with the 1000 Genomes reference population samples and identified 6, 12, 64, and 69 of the individuals with East Asian, African, Admixed American, and European ancestry, respectively^14^. To detect sample swaps, we used NGSCheckMate^17^ and verifyBamID^18^ to call variants from RNA-seq data and assign the best matches between the RNA-seq and genotype data^19^. This led to the removal of 17 samples, leaving 134 samples out of the original 151.

### Proteomics profiling of secretome

#### Sample concentration

We concentrated secretome samples using Amicon® Ultra Centrifugal Filters (2mL, 3kD MWCO, EMD Millipore Corp., Billerica, MA), according to the manufacturer’s instructions, resulting in approximately 60µl of concentrated protein sample after filtering. We measured the protein concentration for a selection of 11 samples using the Bradford assay, which ranged from 0.3µg to 16µg, with an average of approximately 10µg. The total amount of concentrated sample was subsequently used for in-solution digestion.

#### In solution digest

We denatured the concentrated protein samples with 6 mol/L urea, 2 mol/L thiourea, and reduced them using 10 mmol/L dithiothreitol, followed by incubation at 37 °C for 1 hour at 240 rpm. We then cooled the denatured/reduced protein samples to room temperature before alkylating them with 50 mmol/L iodoacetamide in the dark for 1 hour. We precipitated the samples overnight at −20 °C using pre-chilled acetone (5x volume). After centrifuging at 16000 x g for 40 minutes at 0 °C, we subsequently discarded the supernatant. Protein pellets were vacuum centrifuged using a Speed Vac (Thermo Scientific, Savant SPD131DDA), re-suspended in 0.1 mol/L triethylammonium bicarbonate buffer (pH 8.2) containing Trypsin/LysC (1:25 protease:protein, Thermo Scientific), and digested overnight at 37 °C, 240 rpm. We stopped the digestion by acidifying the samples with a final concentration of 1% trifluoroacetic acid (TFA).

#### Peptide purification

We purified the peptide samples using a Bravo AssayMAP liquid handling system (Agilent). After equilibration, we loaded acidified peptide solutions onto AssayMAP C18 Cartridges (Agilent, 5190-6532), washed them using 1% acetonitrile (ACN), 0.1% TFA (aq), and eluted them using 70% acetonitrile, 0.1% TFA (aq). We then vacuum centrifuged the eluted peptides and resuspended them in 20 µl of 2% acetonitrile, 0.05% TFA (aq).

#### LC-MS/MS

We analyzed the samples using untargeted proteomics on a nanoflow LC system (Dionex UltiMate 3000 RSLC nano). We injected the samples onto a nano-trap column (Acclaim® PepMap100 C18 Trap, 5mm x 300um, 5um, 100 Å), at a flow rate of 25 µL/min for 3 mins, using 2% acetonitrile, 0.1% formic acid (FA) in H2O. We then ran the following nano LC gradient at 0.25 µL/min to separate the peptides: 0−5 min, 4-10% B; 5−75 min, 10-30% B; 75-80 min, 30−40% B; 80-85 min, 99% B, 90-120 min 4% B, where A=0.1% FA in H2O, B=80% acetonitrile, 0.1% FA in H2O. The nanocolumn (EASY-Spray PepMap® RSLC C18, 2 μm 100 Å, 75 µm x 50 cm, Cat. No. 160454), set at 45°C, was connected to an EASY-Spray ion source (Thermo Scientific). We collected spectra from an Orbitrap mass analyzer (Q Exactive Plus, Thermo Fisher Scientific) using full MS mode (resolution of 60,000 at 200 m/z) over the mass-to-charge (m/z) range 350–1600. We performed data-dependent MS2 scan using the top 15 ions in each full MS scan (resolution of 15,000 at 200 m/z) with dynamic exclusion enabled.

#### Database Search

We used Thermo Scientific Proteome Discoverer software (version 2.4.0.305) to search raw data files against the human and bovine databases (UniProtKB/Swiss-Prot version 2019_01, 20,413/6,004 protein entries, respectively) using Mascot (version 2.6.0, Matrix Science). We set the mass tolerance at 10 ppm for precursor ions and 20 mmu for fragment ions. We used trypsin as the enzyme with allowance for up to two missed cleavages. We chose carbamidomethylation of cysteine as a fixed modification, and oxidation of methionine, proline, and lysine as variable modifications. We used unique and razor peptides for quantification and exported non-normalized abundance data for further data processing.

#### Data processing

We assessed bovine serum albumin (BSA) contribution to each sample using abundance values. We deemed 11 samples with significantly high levels of BSA to be suppressing mass spectrometry signals of other proteins so they were considered to be outliers and excluded, leaving 123 samples for our study. We filtered protein identifications assigned for both human and bovine species according to razor peptide assignment. Razor peptides are peptides found in multiple protein groups. They are assigned to the protein group with more identified peptides or higher-scoring peptides in case of a tie. This ensures that each instance of spectral evidence is only used for one protein group’s score, avoiding confirmation for multiple groups^20^. We removed proteins with more razor peptides and more matched spectra for Bos Taurus. Proteins filtered out based on this criterion were mostly serum proteins. Using only human proteins, we normalized protein abundances based on the total protein abundance per sample. We scaled the relative quantities of the proteins using log2 transformation.

Additionally, we filtered the dataset to keep only human proteins with less than 30% missing values. After filtering, 1033 human proteins remained, and we imputed their missing values using the KNN-Impute method with k equal to 3 (default value)^21^. We assigned functional categories using Matrisome DB^22^ (http://matrisomeproject.mit.edu/) followed by in-house selection and functional assignment, resulting in a total of 270 ECM and related proteins. Of note, our in-house database of ECM proteins was constructed by extending the Matrisome DB definition of human ECM proteins with additional secreted or extracellular proteins (eg. Apolipoproteins and potentially secreted proteins as predicted by the signalP tool^23^).

#### *Cis*-pQTL and sex-biased pQTL identification

For *cis*-pQTL discovery, we corrected the inverse normal protein abundance data for technical artifacts and unknown technical confounders using the probabilistic estimation of expression residuals (PEER) framework^24^. To optimize for *cis*-pQTL discovery, we performed pQTL mapping using inverse normalized protein abundance residuals corrected with 2, 4, 6, 8, 10, and 12 PEER factors along with sex and 4 genotype principal components (PCs) using tensorQTL permutation pass analysis^25^. We implemented tensorQTL permutation testing to detect the top nominal associated SNP within 1 MB of the transcription start site of a gene, defined as the *cis* region, and with a beta approximation to model the permutation result and correct for all SNPs in linkage disequilibrium (LD) with the most significant SNP (referred here as eSNP) per protein. We used the ‘--permute 1000 10000’ option in tensorQTL. Beta approximated permutation *P*-values were then corrected for multiple testing using the q-value false discovery rate FDR correction^26^. A *cis*-pQTL (eProtein) was defined by having an FDR q-value ≤ 0.20. We report the results of pQTL mapping for 8 PEER factors since we discovered the maximum number of *cis*-pQTL proteins with that many PEER factors.

To determine *cis*-pQTL SNPs with statistically significant different effects on SMC protein abundance in males and females, we conducted a sex-biased *cis*-pQTL analysis using autosomal chromosomes. We used a linear regression model including genotype, 4 genotype PCs, and 8 PEER factors using tensorQTL to test for the significance of genotype-by-sex interaction on protein abundance, since the proportion of male and female sample size was imbalanced. We applied eigenMT, a permutation method, that estimates the effective number of independent tests based on the local LD structure^27^. We considered a sex-biased *cis*-pQTL significant if the eigenMT value <0.05. To classify the identified sex-biased *cis*-pQTLs, we conducted independent linear regression models for each sex-biased pQTL gene and eSNP pair, controlling for four genotype PCs and 8 PEER factors. We calculated the 90% confidence intervals of the effect size (β) for each sex per condition. We considered an effect as positive if the confidence interval did not include 0 and the z-score was positive and an effect as negative, if the confidence interval did not include 0 and the z-score was negative. We classified a sex-biased pQTLs as **1)** sex-biased direction if the confidence intervals differed in sign, **2)** sex-biased magnitude if the confidence intervals were the same sign, and **3)** sex-biased effect if one of the confidence intervals included 0.

### Colocalization between molecular SMC QTLs and vascular disease-related GWAS

We examined whether each *cis*-pQTL is colocalized with vascular disease-related GWAS loci using two different methods. First, we used eQTL and GWAS CAusal Variants Identification in Associated Regions (eCAVIAR)^28^, to test for candidate causal SNPs between our pQTL data and the vascular disease-related GWAS considering variants within a 500 kb window around the eSNP for each eProtein. The maximum number of causal variants was set to 2 and variants were considered colocalized if the colocalization posterior probability (CLPP) was greater than 0.01. We used the 1000 Genomes European reference panel to account for linkage disequilibrium. Finally, we ran Bayesian colocalization analysis using Bayes Factors (COLOC) using the R package COLOC^29^. We input the loci into COLOC with the default priors (p1/p2 = 1 x 10^-4^, and p12 = 1 x 10^-5^), and considered a locus colocalized if PPH4, the hypothesis of a single shared causal variant for both traits within a window, was greater than 0.50. We then plotted and visually inspected all analyzed loci using LocusCompare^30^. Loci that passed both visual inspection and colocalization criteria were considered colocalized.

### SMC pQTL loci in atherosclerosis-related phenotypes

Using the same cohort of primary vascular SMCs from the 151 heart transplant donors, we previously conducted RNA sequencing^31^ and evaluated these cells for three key atherosclerosis-related phenotypes^14^: migration, proliferation, and calcification. We measured calcification in media containing high inorganic phosphate or osteogenic stimuli, while proliferation was measured in control media and media containing PDGF-BB, TGF-β1, or IL-1β. We assessed migration using a modified Boyden chamber assay in a PDGF-BB gradient. We used the Illumina TruSeq mRNA library prep kit to prepare sequencing libraries from SMCs of the 151 donors, and sequenced them to ∼100 million read depth with 300 bp paired-end reads mapped to the hg38 using the STAR Aligner. We found 18,078 expressed genes (> 6 reads/million in at least 80% of the samples). Finally, we identified whether the pQTL loci genes are associated with atherosclerosis-relevant phenotypes.

### SMC cis-pQTL loci in atherosclerotic lesions using single-nucleus ATAC-Seq, scRNA-Seq and gene expression data

We studied the chromatin patterns around pQTLs genes in human atherosclerotic plaque cells using our previously published single-nucleus ATAC-seq data^32^, which included five cell types: endothelial cells (ECs), SMCs, macrophages, natural killer/T cells, and B cells. Additionally, we examined the role of SMC pQTL genes in CAD by analyzing previously published scRNA-seq data from mouse microdissected atherosclerotic lesions from mouse lineage tracing aortic tissues^1^ and human coronary atherosclerotic plaques^33^.

Lastly, we interrogated the gene expression data from 1) human stable and unstable atherosclerotic plaques of carotid arteries (GEO accession number GSE120521)^34^ and 2) early and advanced human atherosclerotic lesions of carotid arteries (GSE28829)^35^.

### Gene silencing, migration, proliferation and calcification

We transfected human aortic SMCs with control (ThermoFisher, 4390846) or LTBP1 (AM16708) siRNA using Lipofectamine RNAiMAX (ThermoFisher, 13778150) per manufacturer’s standard protocol. We collected the cells after 48 hours to measure the extent of downregulation. We extracted the RNA using QIAGEN RNeasy Plus Mini Kit and performed qPCR for *B2M* and *LTBP1* and ECM and SMC genes (*COL1A1, COL1A2, COL11A1, ACTA2, CNN1, TAGLN*) using the primer pairs presented in **Supplementary Table 1**. We used ΔΔCt method to calculate the relative gene expression of *LTBP1* and ECM and SMC genes (COL1A1, COL1A2, COL11A1, ACTA2, CNN1, TAGLN) compared to *B2M* housekeeping gene. Forty-two hours post-transfection, we performed the proliferation assay in a 24-well plate using CCK-8 assays (Dojindo Molecular Technologies, NC9864731) according to the manufacturer’s standard protocol. We performed migration after 24 hours post-transfection with the xCELLigence Biosensor System using specifically designed 16-well plates equipped with membranes with 8-m pores (CIM-plate 16; Roche Diagnostics) as previously described^14^. We seeded the transfected cells in a serum-free medium in the upper chambers and added the chemoattractant PDGF-BB (100 ng/mL) to the lower chambers, with the serum-free medium being the negative control. Cell migration was monitored over 24 h. We analyzed the data using RTCA software version 1.2 (Acea Biosciences Inc., San Diego, CA) combined with R software.

To induce calcification, we used 2.6 mM of inorganic phosphate (Pi). We kept cells with the calcification media for 12 days while changing the media 2-3 times per week. The media without Pi were used as control. At the end of the experiment, we rinsed the wells twice with PBS without Ca^2+^ and Mg^2+^ and then added 50µl 0.6N HCl per well overnight at 4°C. We transferred the supernatant and detached cells from each well into Eppendorf tubes and centrifuged them for 20min at 4°C and 15000g to determine their calcium content using the calcium measurement kit (Randox, CA590) as we previously described^14^. In addition, we collected the cells to measure the extent of downregulation of *LTBP1*, ECM, and SMC genes, as described above.

### Immunohistochemistry staining of LTBP1 in atherosclerotic lesions

#### Mouse atherosclerotic lesions

The University of Virginia Animal Care and Use Committee guidelines and international guidelines were followed for all animal experiments, and we maintained and handled the animals accordingly. We serially sectioned paraffin-embedded brachiocephalic arteries (BCAs) with advanced atherosclerotic lesions at a 10-μm thickness, starting from the aortic arch to the right subclavian artery, in two different mouse lines previously described - SMC-specific conditional knock-out of SMC-Klf4 ApoE-/- KO mice or SMC-Klf4 WT ApoE-/- mice littermate controls^36^. SMC-Klf4 KO lesions were 50% smaller and exhibited features associated with increased plaque stability, including a two-fold increase in the extracellular matrix (ECM) rich α-SMA+ fibrous cap thickness ^36^. To analyze LTBP1-positive SMCs within the BCA, we analyzed sections from a location 750 μm from the start of the branch of the BCA from the aortic arch. We de-paraffinized and rehydrated the BCA sections in xylene and ethanol. After antigen retrieval (H-3300, Vector Laboratories), we blocked the sections with PBS containing fish skin gelatin (6 g/L) and 10% horse serum for 1 hour at room temperature. We incubated the slides with the following antibodies: eYFP detection involved a goat polyclonal anti-GFP antibody (4 μg/mL, ab6673, Abcam) (Overnight 40C), and LTBP1 was detected using a rabbit polyclonal antibody (10 μg/mL, 26855-1-AP, Proteintech) (Overnight 40C). SM α-actin-Cy3 (ACTA2), a mouse monoclonal antibody (4.4 μg/mL, clone 1A4, Sigma Aldrich, C6198), was incubated for 1 hour at room temperature. We used secondary antibodies, including Alexa 488-conjugated donkey anti-goat (A11055, 5 μg/mL, Invitrogen) and Alexa 647-conjugated donkey anti-rabbit (A31573, 5 μg/mL, Invitrogen). For nuclear counterstaining, we used DAPI (0.05 mg/mL, D3571, Thermo Fisher Scientific). We mounted the slides using Prolong Gold Antifade (Invitrogen, P36930). We imaged the immunofluorescent staining using a Zeiss LSM880 confocal microscope to acquire a series of z-stack images at 1-μm intervals. We analyzed each z-stack image using Zen 2009 Light Edition Software (Zeiss) and performed single-cell counting to phenotype and quantify the cell population within the 30μm thick layer proximal to the lumen (fibrous cap area).

#### Human atherosclerotic lesions

The Institutional Review Board at CVPath Institute (Gaithersburg, Maryland) approved this study. We selected five human coronary artery specimens with advanced atherosclerotic lesions of thin- and thick-cap fibroatheroma from the CVPath Institute Sudden Death Registry. The artery segments were soaked in formalin and embedded 2 to 3-mm segments in paraffin. From each segment, we cut cross-sections that were 5-8 microns thick and mounted them on charged glass slides. We performed staining with hematoxylin-eosin (H&E) and Movat Pentachrome. The H&E and Movat Pentachrome stained slides were developed using the labeled streptavidin-biotin (LSAB) and NovaRed kit (Vector Laboratories). Images were captured using a 10X or 20X objective on the Axio Scan Z1 microscope (Zeiss, Germany), and staining images were visualized using Zen (Zeiss, Germany). Immunohistochemistry for LTBP1 and smooth muscle actin (SMA) was carried out on 5-8 microns thick sections. We incubated the slides overnight with the LTBP1 antibody (1:50 dilution, 26855-1-AP, Proteintech) and developed using a 1:150 dilution of donkey anti-rabbit Alexa fluor 555 secondary antibody (A31572, Invitrogen) for 2 hours followed by 1 hour with SMA (ACTA2/FITC conjugated) antibody (1:500 dilution, F3777 Sigma). The slides were imaged using either a LSM 980 or LSM 800 confocal microscope at 20-40X resolution (Zeiss, Germany).

## RESULTS

### Proteomics profiling of human aortic SMCs secretome

We performed proteomic profiling of collected cell culture media from primary aortic SMCs derived from 151 ancestrally-diverse healthy heart transplant donors. After quality control filtering, we performed analyses on 123 donors (100 males and 23 females) (**Figure 1**). We reliably detected the abundance of 270 secreted ECM proteins (**Supplementary Table 2**). Among these, fibrillar collagens, fibronectin, basement membrane proteins, Secreted Protein Acidic And Cysteine Rich (SPARC) protein, and small leucine-rich proteoglycans, are all involved in vascular diseases^37–40^. The number of secreted and ECM-related proteins quantified in the current study was lower than the number of unique genes encoding for ECM proteins from the same donors in our previous study^14^ (270 *vs*. 464), possibly due to lower sensitivity of the LC-MS/MS approach. However, for the majority of the proteins, we found a strong correlation between their abundance and the expression level of the transcripts of the same genes as measured by RNA sequencing^19^ (**Supplementary** Figure 1A**, Supplementary Table 3**). The number of significant positive correlations (89%) was much higher than significant negative correlations (3%). In contrast, 8% of the protein-mRNA pairs was not correlated (**Supplementary** Figure 1B).

**Figure 1:**
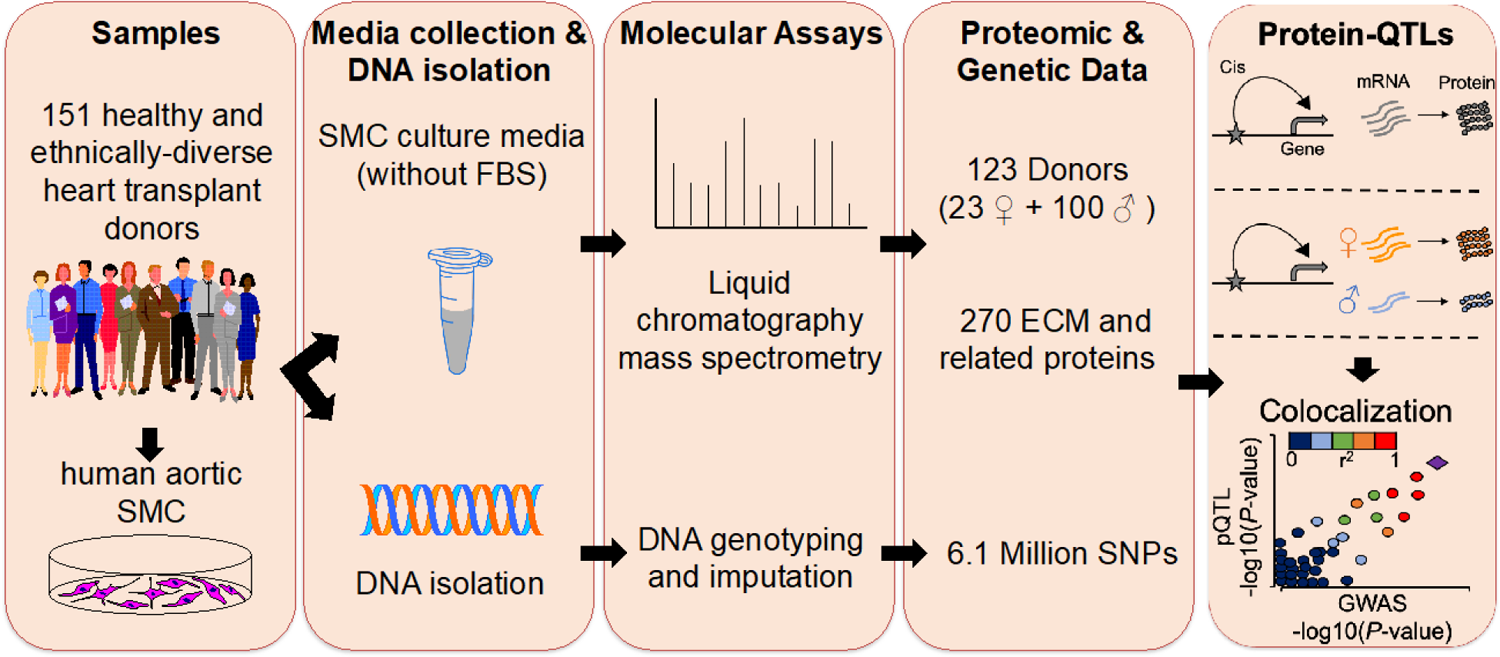
Study design and overview of analyses. We cultured aortic smooth muscle cells (SMCs) from 151 multi-ancestry heart transplant donors in 5% fetal bovine serum (FBS) media until 90% confluence. We then replaced the cell cultures with new media without FBS. We collected the media after 24 hours for proteomics profiling. In parallel, the DNA was isolated and subjected to genotyping and imputation. We calculated the associations of proteins with the genotypes of ∼6.1 million imputed SNPs to discover *cis*-protein quantitative trait loci (pQTLs) and sex-biased pQTLs. We identified the colocalization between molecular QTL and vascular disease-related GWAS loci associations using two different methods. ECM: extracellular matrix, Protein-QTLs: Protein quantitative trait locus.

### *cis*-protein Quantitative Trait Loci in SMCs

To identify genetic loci associated with protein abundance, we conducted association mapping with the genotypes of ∼6.1 million variants and the abundance of 270 secreted proteins using tensorQTL^25^. We identified MMP1 with a *cis*-pQTL (< 1Mb) at FDR q-value ≤ 0.05 (Corresponding *P*-value ≤ 8.1×10^-8^). When we relaxed the significance threshold to 20% FDR q-value, we identified 19 additional proteins. (**Table 1, Supplementary Table 4**). Conditioning on the lead SNPs of the 20 proteins did not identify secondary pQTLs. All the identified pQTL variants were associated with the expression of a single protein. Using our previously-published expression QTL (eQTL) results^19^, we identified SNPs associated with both mRNA and protein abundance. We used two different colocalization approaches, eCAVIAR and COLOC^26,25^, and identified 4 loci that were associated with both the expression and protein abundance of LAMA5, LTBP1, MMP1, and LAMC2 (**Figures 2 and 5**). As expected, we found significantly high correlations between the protein and transcripts abundance for these 4 genes with concordant QTL compared to the genes with only eQTL or pQTL (**Supplementary Table 3)**.

**Figure 2:**
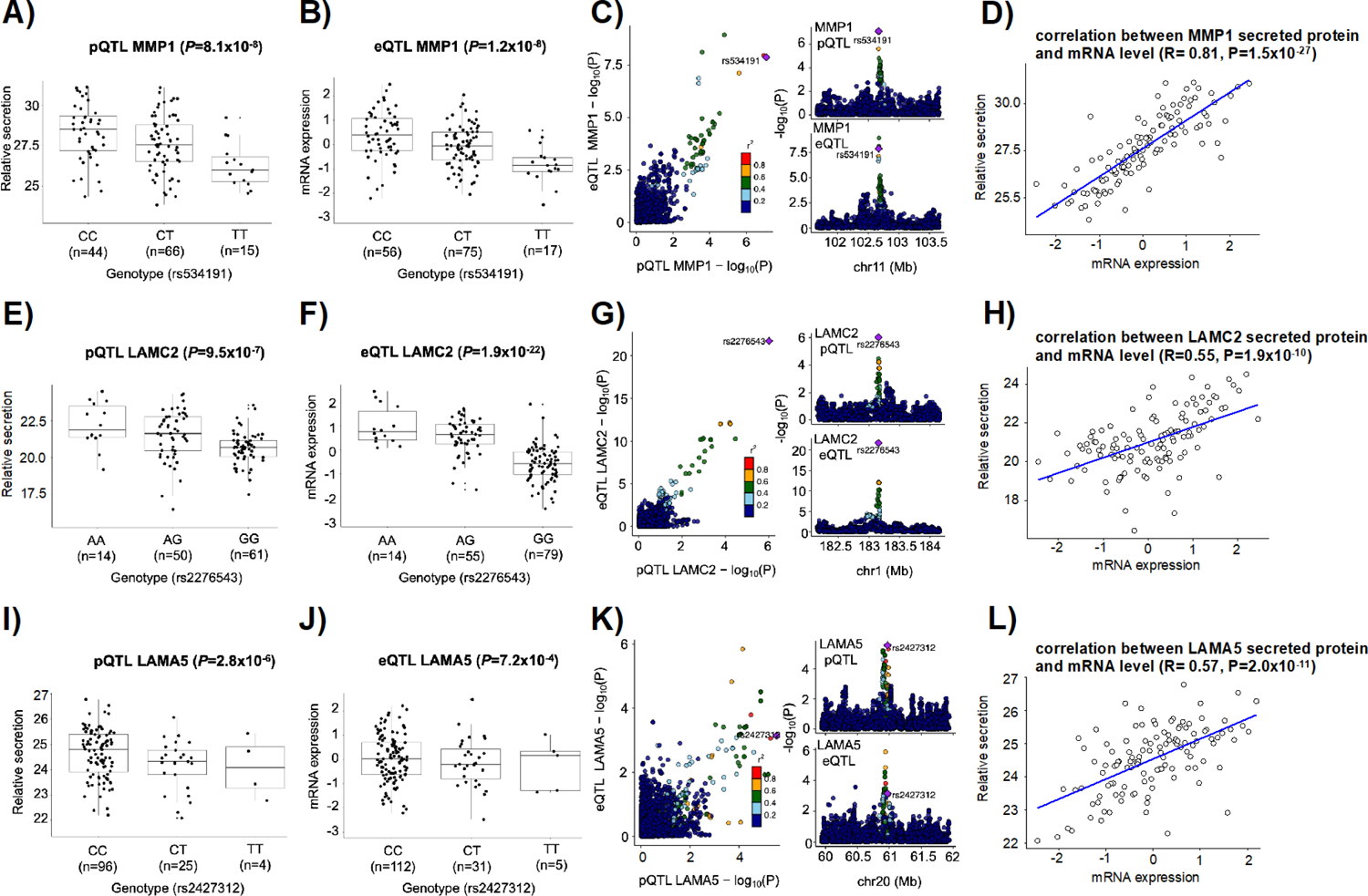
Genotype-phenotype and regional association plots of the colocalized eQTLs and pQTLs. Box and whisker plots of the associated secreted SMC protein (**A, D, G**) and transcript (**B, E, H**) in relation to the genotype of the three lead pQTL variants. See figure 6 A-C for the fourth regional association plots of the colocalized eQTLs and pQTLs. *P*-values were determined using the linear mixed-model regression in performing eQTL or pQTL mapping. Associations with genetic variants and protein and expression levels around the lead SNP for each colocalized locus are shown using LocusCompare (**C, F, I**). Linkage disequilibrium (r^2^) of each SNP with the lead SNP is color-coded based on European ancestry populations in the 1000 Genomes project. eQTL: Expression quantitative trait locus, pQTL: Protein quantitative trait locus.

**Table 1:** Protein quantitative trait loci identified for secreted aortic smooth muscle cell proteins.

| Protein ID | SNP ID | Chromosome | Position (hg38) | $\beta$ | $\beta(\text{se})$ | qvalue |
| --- | --- | --- | --- | --- | --- | --- |
| MMP1 | rs534191 | chr11 | 102800966 | -0.564 | 0.098 | 3.06E-02 |
| LAMC2 | rs2276543 | chr1 | 183186170 | 0.598 | 0.115 | 8.30E-02 |
| LTBP1 | rs6739323 | chr2 | 33163275 | -0.392 | 0.075 | 8.30E-02 |
| ITGBL1 | rs2152321 | chr13 | 101485015 | 0.655 | 0.128 | 9.62E-02 |
| NPC2 | rs118047935 | chr14 | 74268459 | -0.708 | 0.141 | 9.62E-02 |
| ITIH2 | rs10905183 | chr10 | 7565197 | 0.343 | 0.070 | 1.20E-01 |
| HSPB6 | rs12460459 | chr19 | 35088257 | 0.934 | 0.197 | 1.20E-01 |
| ITGAV | rs12988401 | chr2 | 186798551 | 0.497 | 0.110 | 1.20E-01 |
| CST3 | rs6048357 | chr20 | 22800518 | -0.792 | 0.168 | 1.20E-01 |
| LAMA5 | rs2427312 | chr20 | 62395535 | -0.654 | 0.132 | 1.20E-01 |
| FBLN2 | rs111338444 | chr3 | 14013315 | -0.662 | 0.136 | 1.20E-01 |
| FSTL1 | rs28600057 | chr3 | 120129510 | -0.531 | 0.116 | 1.20E-01 |
| HAPLN1 | rs13188822 | chr5 | 83925733 | 0.523 | 0.110 | 1.20E-01 |
| ECM2 | rs10820900 | chr9 | 91733326 | 0.545 | 0.118 | 1.20E-01 |
| LAMA2 | rs7738133 | chr6 | 129080895 | 0.575 | 0.125 | 1.34E-01 |
| CTSB | rs55894533 | chr8 | 11891733 | -0.473 | 0.102 | 1.36E-01 |
| LOXL3 | rs1987965 | chr2 | 75389537 | -0.442 | 0.097 | 1.37E-01 |
| LGALS3BP | rs7218341 | chr17 | 78259697 | 1.051 | 0.222 | 1.50E-01 |
| HSP90AB1 | rs77294443 | chr6 | 44615088 | 0.957 | 0.210 | 1.50E-01 |
| COL13A1 | rs7090788 | chr10 | 69453894 | 0.596 | 0.131 | 2.00E-01 |

Since sex differences in the genetic regulation of gene expression have been detected in many tissues^19,41,42^, we separately identified sex-biased *cis*-pQTLs in SMCs. We identified three sex-biased *cis*-pQTLs (eigenMT value <0.05) for COL5A1, ABI3BP and SOD3. (**Supplementary figure 2, Supplementary Table 5**).

### Colocalization between pQTLs and vascular-related diseases GWAS signals

To test whether secreted proteins by SMCs contribute to the genetic risk for vascular-related diseases involving SMCs, we identified the common genetic signals for SMC pQTL loci with vascular disease-related GWAS loci using two different approaches: eCAVIAR and COLOC^28,29^. We downloaded the summary statistics for CAD, myocardial infarction, stroke, aortic aneurysm, and blood pressure GWAS from the GWAS catalog and the UK Biobank^43^ (**Supplementary Table 6**). We used colocalization posterior probability (CLPP) > 0.01 as cutoffs for eCAVIAR and PPH4 > 0.5 for COLOC. We identified one blood pressure and one pleiotropic CAD/ aortic aneurysm locus associated with secreted SMC protein abundance. These loci were associated with the abundance of Hornerin (HRNR) and Laminin Subunit Gamma 2 (LAMC2) proteins (**Figure 3, Supplementary Table 7-8**). Since the two proteins predicted by the distinct colocalization methods differed, we visualized the coincidence of the pQTL and GWAS lead SNPs by inspecting the regional colocalization plots using LocusCompare^30^. This coincidence was also supported by conditional analysis on each lead SNP and GWAS index SNP. The CAD and aortic aneurysm risk allele (C) of SNP rs6681093 is associated with lower HRNR protein secretion from SMCs (**Figure 3C)**. In contrast, the diastolic blood pressure risk allele (T) of SNP rs10797858 is associated with lower LAMC2 protein secretion from SMCs (**Figure 3E)**.

**Figure 3:**
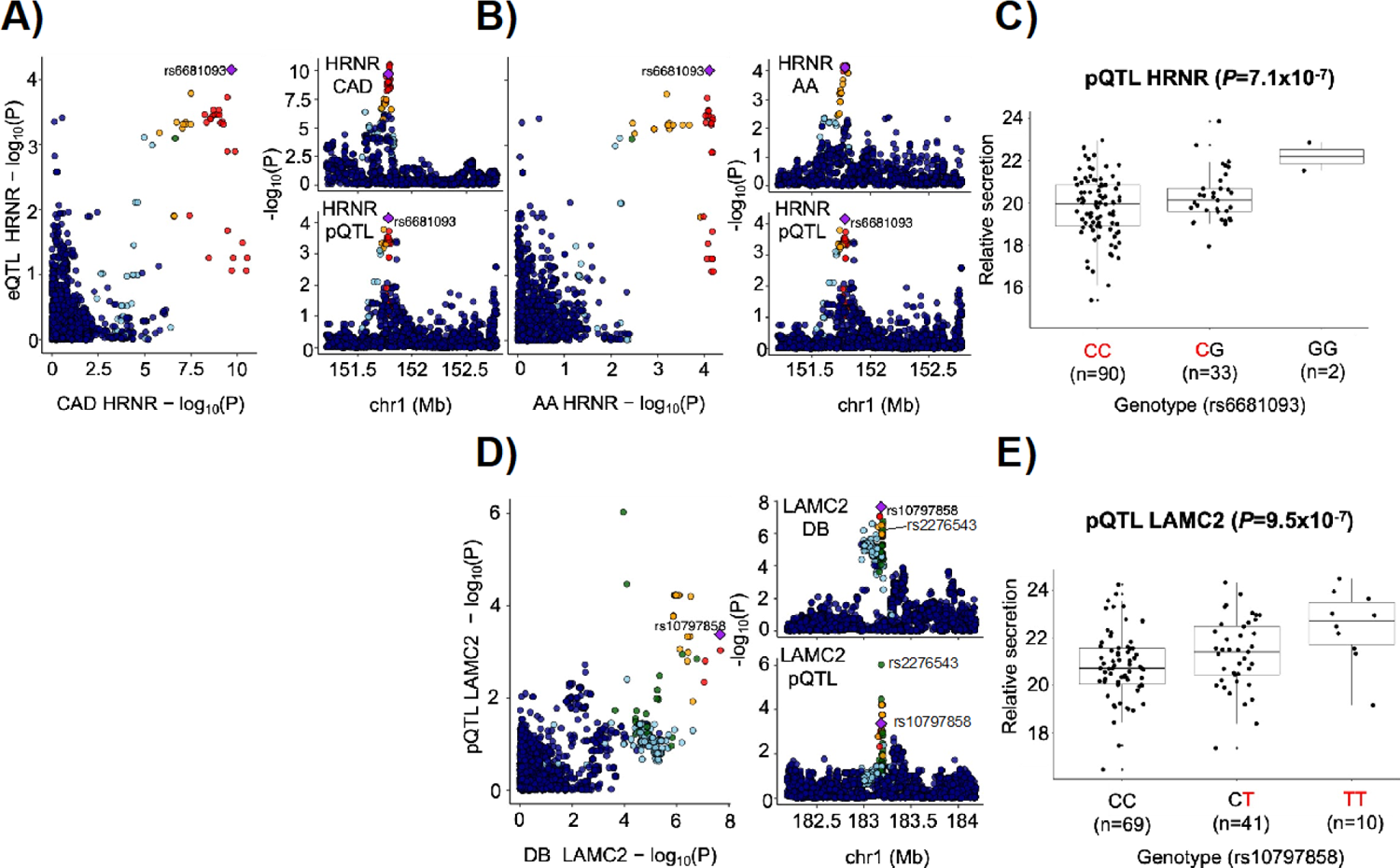
Colocalization between pQTLs and vascular disease-related GWAS signals. *Cis*-pQTL for Homerin (HRNR) protein secretion colocalized with the 1q21.3 **A)** coronary artery disease (CAD) and **B)** aortic aneurysm (AA) GWAS loci. **C)** The risk allele (C) of SNP rs6681093 is associated with lower HRNR protein secretion from SMCs. *Cis*-pQTL for Laminin Subunit Gamma 2 (LAMC2) protein secretion colocalized with the 1q25.3 **D)** Diastolic blood pressure (DBP) GWAS locus. **E)** The risk allele (T) of SNP rs10797858 is associated with lower LAMC2 protein secretion from SMCs.

### Functional annotation of SMC pQTLs

To provide additional support for the involvement of secreted proteins with pQTLs in vascular diseases, we conducted lookups in published datasets (**Supplementary** Figure 3). First, we interrogated the gene expression data from 1) human stable and unstable atherosclerotic plaques of carotid arteries (GEO accession number GSE120521)^34^ and 2) early and advanced human atherosclerotic lesions of carotid arteries (GSE28829)^35^. Of the 20 identified proteins with *cis*-pQTL, 10 were significantly altered at the transcript level in one of the comparisons: Cathepsin B (CTSB), Matrix metallopeptidase 1 (MMP1), Transforming growth factor beta binding protein 1 (LTBP1), Niemann-Pick disease type C2 (NPC2), Heat shock protein family B (Small) member 6 (HSPB6), Laminin subunit alpha 5 (LAMA5), Follistatin-like 1 (FSTL1), Extracellular matrix protein 2 (ECM2), Heat shock protein 90 alpha family class B member 1 (HSP90AB1) and Collagen type XIII alpha 1 chain (COL13A1) (**Figure 4, Supplementary Table 9**). LTBP1, HSPB6, LAMA5, FSTL1, ECM2, HSP90AB1, and COL13A1 were upregulated in stable or early lesions compared to unstable or advanced lesions. While CTSB, MMP1, and NPC2 were downregulated in at least one of the stable or early lesions compared to unstable or advanced lesions.

**Figure 4:**
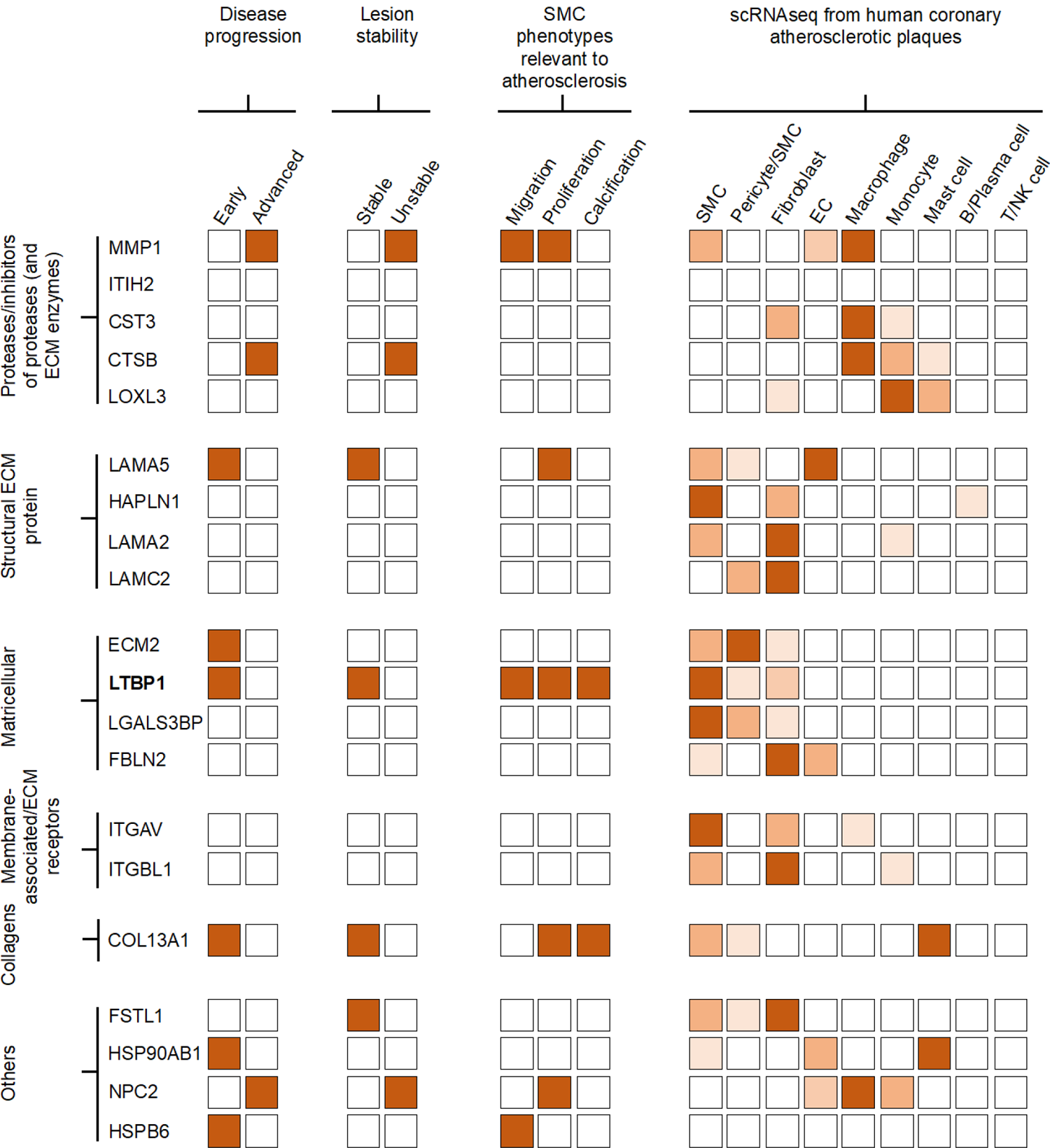
Overview of the multi-omics data used to functionally annotate the 20 pQTL proteins. Orange color indicates either the genes encoding proteins with *cis* pQTLs are upregulated in different stages^34^ of disease progression and lesion stability^33^, or significantly correlated with SMC phenotypes relevant to atherosclerosis or highly expressed in single cells isolated from human coronary atherosclerotic plaques^13^. For scRNAseq data, we display the normalized expression values (TPM) range above 0 to +1.5. Dark orange color indicates higher expression, while light color indicates lower expression.

In a second approach, to identify which of the 10 pQTL loci harboring LTBP1, HSPB6, LAMA5, FSTL1, ECM2, CTSB, HSP90AB1, COL13A1, MMP1, and NPC2 might be associated with an atherosclerosis-relevant phenotype in SMCs, we interrogated our previously published data on quantifying 12 atherosclerosis-relevant phenotypes in the same donors^14^. LTBP1, LAMA5, HSBP6, COL13A1, NPC2, and MMP1 protein levels showed significant correlations with migration, proliferation, or calcification of SMCs. (**Figure 4, Supplementary Table 10**).

Third, we interrogated single-cell RNAseq (scRNAseq) data from human coronary atherosclerotic plaques and confirmed the expression of the genes for 19 of the 20 proteins with *cis*-pQTLs in the SMC clusters (**Figure 5, Supplementary** Figure 4)^33^. 10 of the 19 genes had higher expression in SMCs, pericytes, and fibroblasts compared to endothelial cells, monocytes, macrophages, and other immune cells.

**Figure 5:**
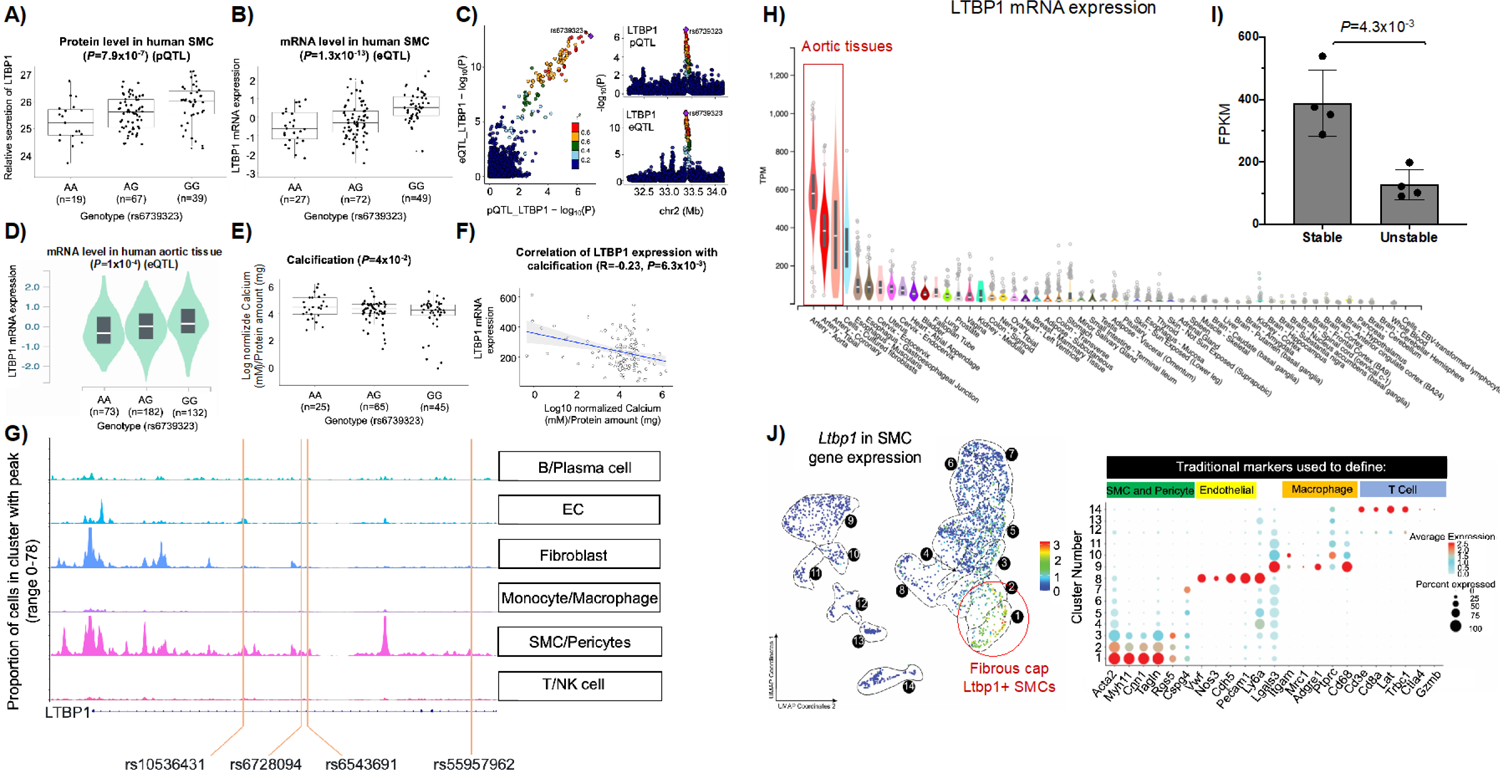
Molecular and cellular characterization of the *LTBP1* locus. The rs6739323-A variant is associated with lower abundance of **A)** secreted *LTBP1* and **B)** mRNA in human SMCs. **C)** Colocalization analyses between pQTL and eQTLs predicted rs6739323 as the causal variant affecting *LTBP1* expression in the 2p22.3 locus. The rs6739323-A variant was also associated with **D)** lower expression of *LTBP1* at the mRNA level in human aortic tissue^30^ and with **E)** higher SMC calcification^11^. **F)** Negative correlation between *LTBP1* expression and SMC calcification was observed. **G)** Single-cell ATACseq analysis from human coronary atherosclerotic plaques revealed the SMC specificity of LTBP1 regulatory elements^71^. SNPs rs10536431, rs6728094, rs6543691, and rs55957962 that are in high LD (r^2^≥0.8) with rs6739323 are located in an accessible chromatin region identified by ATACseq in SMCs (lower panel). **H)** Gene expression of *LTBP1* in human tissues that are part of the GTEx project showed higher expression in aortic tissues^30^. **I)** Gene expression of *LTBP1* in human unstable atherosclerotic plaques of carotid arteries compared to stable lesions^27^. **J)** scRNA-seq analyses in mouse aortic tissues from microdissected atherosclerotic lesions revealed that *Ltbp1* is predominantly expressed in MYH11+ACTA2+ lesion SMCs that are known to play a critical role in maintaining fibrous cap stability^34^.

Our functional annotation approach prioritized the *LTBP1* gene at the chr2q22.3 locus, whose expression is genetically regulated in SMCs (**Figure 4, Supplementary** Figure 3). The rs6739323-A variant was associated with lower expression of LTBP1 at the protein (**Figure 5A**) and mRNA levels (**Figure 5B**). Colocalization analyses between pQTL and eQTLs predicted rs6739323 as the causal variant affecting LTBP1 expression (**Figure 5C**). Using the GTEx data set^44^, we replicated our finding that the rs6739323-A allele was associated with lower expression of LTBP1 at the mRNA level in the aortic tissue (**Figure 5D).** The SNP was not associated with LTBP1 expression in monocytes/macrophages^45^ or aortic endothelial cells^46^. Furthermore, the rs6739323-A variant was associated with higher calcification^47^ in our SMC dataset (**Figure 5E)**. We also observed a negative correlation between *LTBP1* expression and SMC calcification (**Figure 5F**). Human single-cell ATACseq analysis from coronary atherosclerotic plaques revealed the SMC specificity of LTBP1 regulatory elements (**Figure 5G**). Gene expression of *LTBP1* in human tissues that are part of the GTEx project revealed high enrichment of LTBP1 in all three artery tissues: artery aorta, tibial, and coronary (**Figure 5H**). *LTBP1* was potently downregulated in human unstable atherosclerotic plaques of carotid arteries compared to stable lesions (**Figure 5I**). scRNA-seq analyses in mouse lineage tracing aortic tissues^1^ from microdissected atherosclerotic lesions revealed that *Ltbp1* was predominantly expressed in differentiated SMC, including MYH11+ACTA2+ lesion SMCs that are known to play a critical role in maintaining fibrous cap stability (**Figure 5J**). siRNA-mediated LTBP1 downregulation (64% decrease) (**Figure 6A**) after 48 hours led to a 2.4 fold decrease in proliferation (**Figure 6B**), migration (2.8-fold) (**Figure 6C**), an increase in calcification (1.7-fold) of SMCs (**Figure 6D**), and decrease of ECM (**Figure 6E-G**) and SMC (**Figure 6H-J**) marker genes. However, after 12 days of transfection, downregulation of LTBP1 in SMCs results (**Supplementary** Figure 5A and H) in a significant increase in the expression of ECM genes both in non-calcified (**Supplementary** Figure 5B-D) and calcified media (**Supplementary** Figure 5I-K). Additionally, SMC-specific marker genes exhibit increased expression in non-calcified media (**Supplementary** Figure 5E-J) and a more pronounced increase in expression in calcified media (**Supplementary** Figure 5L-N). Collectively, these studies suggest that the rs6739323 variant at the 2q22.3 locus affects atherosclerosis-relevant SMC phenotypes by modulating *LTBP1* expression.

**Figure 6:**
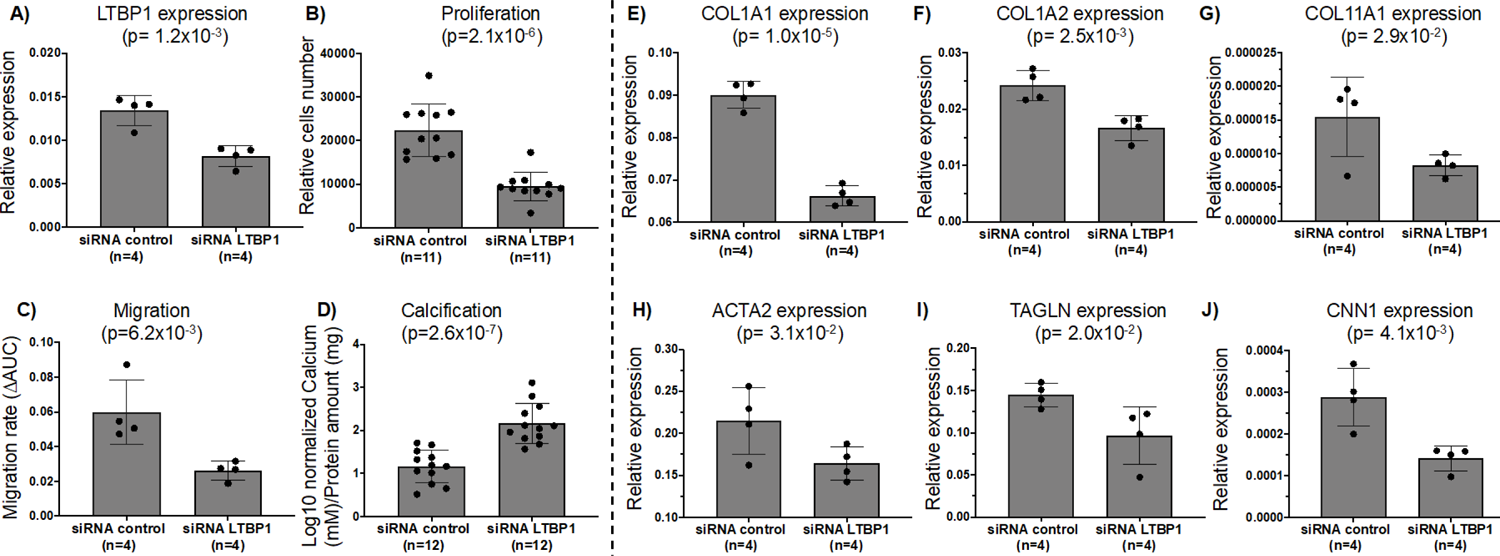
Modulation of *LTBP1* expression in SMCs. **A)** *LTBP1* downregulation in SMCs leads to a significant decrease in **B)** proliferation and **C)** migration, an increase of **D)** calcification and decrease of the expression of **E-G)** ECM genes, and **H-J)** SMC-specific marker genes. n represents the technical repeats from the same donor.

Based on our human genetics and *in vitro* findings, we hypothesized that LTBP1 may play a role in maintaining fibrous cap stability in atherosclerotic lesions. Previous studies have shown that hypercholesterolemic ApoE-null mice lacking KLF4 expression in their SMC exhibit reduced SMC content, decreased fibrous cap thickness, and a two-fold increase in ECM deposition, leading to increased plaque stability compared to wild-type controls ^36^. To test our hypothesis, we quantified the expression of LTBP1 in ACTA2-positive SMCs within stable lesions of hypercholesterolemic SMC-specific conditional knockout (SMC-Klf4 ApoE-/- KO) mice and compared it to SMC-Klf4 wild-type (WT) ApoE-/- control mice. As previously described, these mice were fed a Western diet for 26 weeks, resulting in advanced vulnerable atherosclerotic lesions^36^. Immunofluorescence staining revealed the presence of LTBP1 protein in SMCs of the aorta and fibrous cap (**Figure 7A**). We observed a higher number of ACTA2-positive SMCs expressing LTBP1 in stable plaques of SMC-Klf4 ApoE-/- KO mice compared to SMC-Klf4 WT ApoE-/- control mice (**Figure 7B**). Additionally, the fraction of SMC-derived LTBP1+ cells (Myh11-eYFP+LTBP1+/eYFP) was increased in the fibrous cap of SMC-Klf4 ApoE-/- KO mice (**Figure 7C**). Moreover, the fraction of α-SMA+ cells co-expressing LTBP1 was higher in the fibrous cap of SMC-Klf4 KO mice compared to WT control mice (**Figure 7D**). The fractions of α-SMA+ cells expressing LTBP1, both among all LTBP1+ cells and all (DAPI+) cells, were also increased in the fibrous cap area (**Figure 7E-F**). However, the SMC-derived LTBP1 fraction among all LTBP1+ cells in the fibrous cap remained unchanged (**Figure 7G**).

**Figure 7:**
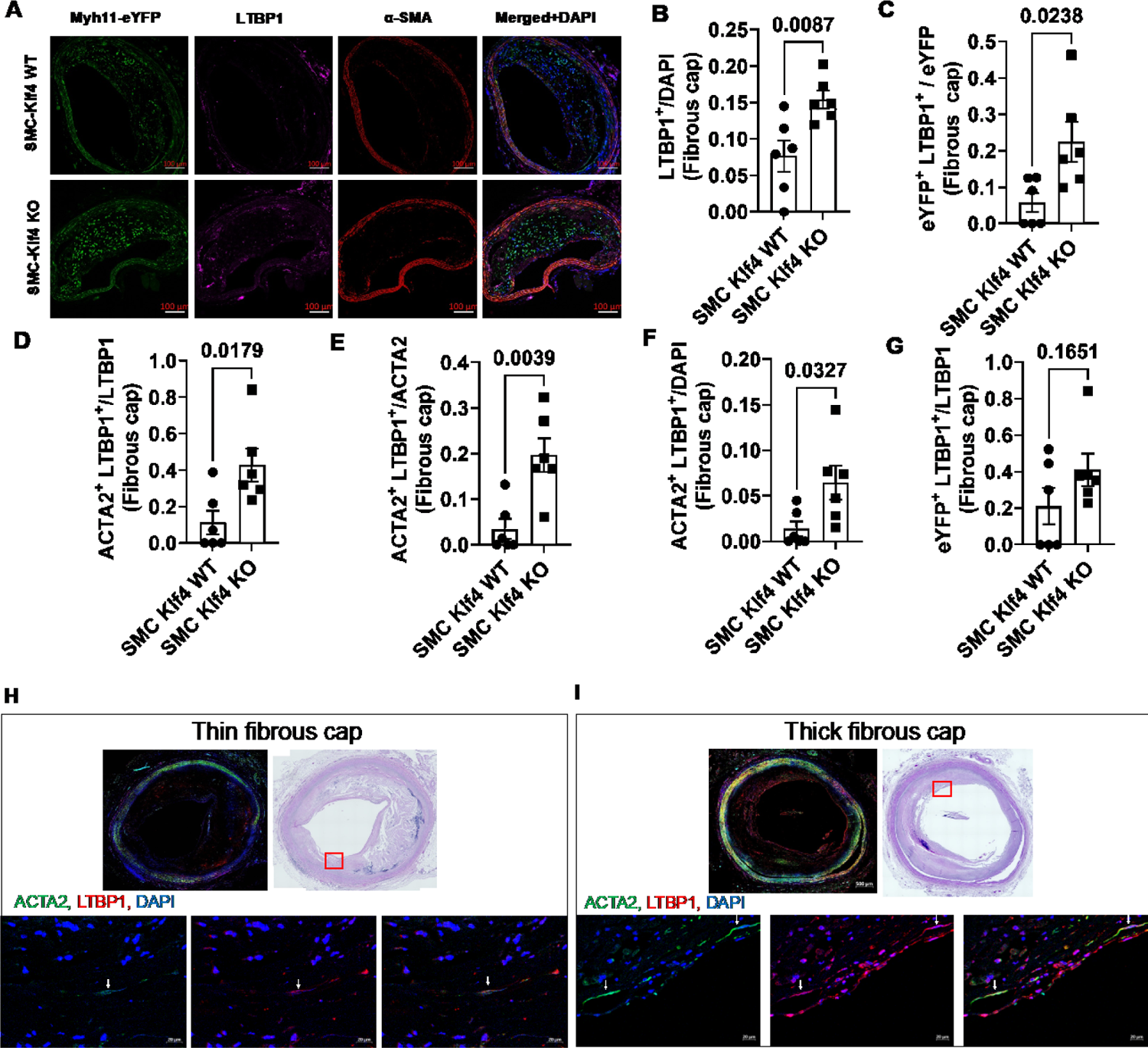
LTBP1 immunostaining in mouse and human atherosclerotic lesions. **A)** Representative immunofluorescence images of brachiocephalic artery (BCA) lesions from SMC-Klf4 ApoE-/- KO and SMC-Klf4 WT ApoE-/- mice, which were fed 26 weeks of hypercholesterolemic Western diet, were stained with eYFP, ACTA2, LTBP1, and DAPI. **B)** LTBP1+ cells of all (DAPI+) cells in the fibrous cap. **C)** Myh11-eYFP+ (SMC)-derived LTBP1+ cells of all Myh11-eYFP+ SMC cells in the fibrous cap. **D)** Fraction of ACTA2+ cells also LTBP1+ of all ACTA2+ cells in the fibrous cap. **E)** Fraction of ACTA2+ cells also LTBP1+ of all LTBP1+ cells in the fibrous cap. **F)** ACTA2+ cells also LTBP1+ cells of all (DAPI) cells in the fibrous cap. **G)** Myh11-eYFP+ (SMC)-derived LTBP1+ cells of all LTBP1+ cells in the fibrous cap. Error bars represent mean ± SEM; p-values displayed refer to Mann-Whitney U test between SMC Klf4 WT and KO groups. Each circle or square shape on the graph indicates data from an individual mouse. Single-channel images with isotype controls are shown in **Supplementary** Figure 6. **H-I)** Results of LTBP1, ACTA2 staining in human coronary artery fibrous cap atheroma. IgG control images are shown in **Supplementary** Figure 7. The representative images in A, H, and I were chosen since they captured the critical lesion features, including the three anatomical layers, cell composition, and evident staining.

We also studied LTBP1 expression in thin and thick fibrous cap regions of human coronary artery lesions. We observed a significant reduction in LTBP1 expression in ACTA2-positive SMCs within the thin-cap region of late-stage fibroatheroma compared to SMCs within the protective thick fibrous cap of human coronary artery lesions (**Figure 7H-I**). This finding suggests that lower LTBP1 expression in SMCs, leading to decreased proliferation and migration and increased calcification, may contribute to plaque vulnerability and increase the risk of coronary artery disease (CAD) due to weakened fibrous caps prone to rupture.

## DISCUSSION

Previous studies have shown that SMCs could have beneficial or detrimental roles in cardiovascular diseases ^1,36,48,49^. In atherosclerotic disease, SMCs overlay the atheroma and secrete ECM components which stabilize the “fibrous cap”, the thickness and composition of which is one of the key determinants of plaque stability. However, they can also give rise to foam cells and osteochondrogenic cells that are detrimental for plaque stability and overall pathogenesis^1,2^. SMC phenotypic changes also play a role in other vascular diseases such as hypertension via changes in contractility and remodeling of resistance arteries, and development of aortic aneurysms where they contribute, along with macrophages, to disruption of the ECM^5^. However, the interaction of SMCs and ECM in vascular diseases is complex and not fully understood. Using our unique collection of human aortic SMCs from 123 healthy heart transplant donors, we performed genome-wide pQTL analyses of the secreted ECM and related proteins. Since we had measured both mRNA and protein abundance in the same cells, we were able to demonstrate a significant correlation between transcript and protein levels. Further, the relationship between our SMC pQTL and eQTL^14^ discoveries revealed that only a small portion of QTL variants was associated with both protein abundance and mRNA expression, in agreement with other studies^50,51^. Multiple reasons could explain this discrepancy between mRNA expression and protein abundance. The unequal number of expressed transcripts and proteins could partly be due to the differences in technological platforms, data analysis approaches, and the sample size used in eQTL (139 samples) and pQTL (123 samples) studies. Lastly, protein levels can be affected by additional post-transcriptional and post-translational regulatory mechanisms in contrast to mRNA levels. These results indicate the importance of studying the gene expression at both the mRNA and protein levels to comprehend the impact of genetic variants on molecular phenotypes. Next, we utilized a systems genetics approach to understand the disease relevance of secreted SMC proteins. Colocalization between molecular SMC QTLs and vascular disease-related loci allows for the prediction of causal genes and SNPs for hitherto unsolved association signals. Our previous eQTL study identified that 58 and 127 CAD loci were explained by changes in gene expression and alternative splicing, respectively^19^. Here, we provide evidence that ECM proteins secreted by SMCs contribute to the genetic architecture of cardiovascular diseases by colocalizing pQTLs with vascular disease-related GWAS loci. We identified HRNR– which colocalizes with CAD and aortic aneurysm, and LAMC2-which colocalizes with blood pressure GWAS loci. Fu et al. showed that HRNR promotes hepatocellular carcinoma tumor progression^52^ and the knockdown of *HRNR* significantly inhibited the proliferation, colony formation, migration and invasion of hepatocellular carcinoma tumor cells^52^. However, the role of HRNR in SMC biology or cardiovascular disease is not yet studied. Similarly, no functional study of LAMC2 has been carried out in the context of vascular biology and cardiovascular diseases; however, several studies showed its involvement in cancer^53–57^ suggesting that LAMC2 plays a role in proliferation. To date, most proteomics studies have been conducted using plasma samples from patients at higher risk for cardiovascular diseases^58–62^. However, cell type-specific proteomic analyses using disease-relevant cells will be a valuable tool to dissect the origin of the proteome to accelerate the pathway to translation.

Identifying a reliable biomarker to distinguish between stable and unstable atherosclerotic plaques has proven to be challenging. Based on immunohistology, it is well-established that atherosclerotic lesions with thick fibrous caps, rich in MYH11+ and ACTA2+ SMCs relative to CD68+ macrophages are more stable and less prone to rupture^36,48,63,64^. In addition, SMCs are the main producers of ECM including collagen and proteoglycans in the fibrous matrix^8^. In the present study, we identified 10 SMC pQTLs that were differentially expressed at mRNA level in stable and unstable human atherosclerotic lesions, possibly acting by modulating ECM synthesis and secretion to affect the disease. These loci harbor LTBP1, HSPB6, LAMA5, FSTL1, ECM2, CTSB, HSP90AB1, COL13A1, MMP1, and NPC2. For example, MMP1, CTSB, LAMA5 and FSTL1 protein levels were genetically regulated in our study. LAMA5 and FSLT1 transcript levels were upregulated in stable lesions and MMP1, CTSB transcript levels were upregulated in unstable lesions. Papaspyridonos et al. previously showed that LAMA5 was upregulated in stable lesions lesions^65^, while MMP1 and CTSB were upregulated in unstable lesions. Langley et al. also showed that CTSB was upregulated in unstable atherosclerotic lesions^66^. Growing evidence indicates that increased protease activity causes plaque rupture, with candidate proteases including cathepsins, and MMPs, including MMP1^67^. Studies also showed that FSTL1 acts as an anti-inflammatory factor that protects against ischemic heart disease^68^. LAMA5 was more enriched in stable human plaque lesions compared to ruptured lesions^69^. These proteins (MMP1, CTSB, LAMA5 and FSTL1) and the remaining 6 are promising biomarkers of plaque stability and instability since they are expressed in the atherosclerotic plaque at the mRNA or protein levels. In addition, these proteins were altered during the disease progression and plaque stability, suggesting that they may be secreted by SMCs in atherosclerotic lesions.

To validate our pQTL findings, we conducted functional annotation of the identified SMC pQTLs. This approach prioritized a genetic variant, rs6739323-A, at the 2p22.3 locus associated with lower expression of *LTBP1*, a gene encoding latent-transforming growth factor beta-binding protein 1, in SMCs and atheroprone areas of the aorta; and increased risk for SMC calcification. Further, LTBP1 expression was decreased in unstable and advanced atherosclerotic plaque lesions. We observed significant decreases in SMC and ECM marker gene expression after 48 hours of LTBP1 downregulation. However, after 12 days, we noticed an increase in the expression of SMC and ECM genes, both under calcified and non-calcified conditions. It is possible that compensatory mechanisms were triggered during the 12-day period of LTBP1 downregulation, which could explain the significant increase in the expression of SMC and ECM genes. LTBP1 is an ECM protein essential for secretion, activation, and extracellular localization of TGF-β^70^. Previous studies have demonstrated that TGF-β plays an atheroprotective role in SMCs^71^. We found that LTBP1 expression is highly enriched in SMCs based on scRNAseq of human arteries^1,33^. Immunohistochemistry studies using mice and rats demonstrated the presence of fibrillar LTBP1 colocalized in the ECM with collagen, fibronectin, and fibrillin^72–74^. Together, our results suggest that LTBP1 may regulate differentiated SMC functions perturbed during atherosclerotic disease. Several studies have linked LTBP1 expression with different types of tumors^75–79^. However, to date, little is known regarding its role in atherosclerosis. Our results imply that LTBP1 could be a prognostic marker for atherosclerosis, and further investigation of the function of this protein in plaque destabilization is merited.

We acknowledge some limitations of our study. For example, the list of identified proteins is not exhaustive for all proteins potentially involved in plaque stability since we focused on the proteins secreted by SMCs, however atherosclerotic plaques have many more distinct cell types^1,32,33^. Moreover, although there is strong evidence that transcript and protein levels of some of the identified genes indeed reflect abundance in the plaque, with SMCs being a potential source, direct support for this concept needs *in vivo* validation. In addition, the technology we used in our study to measure protein concentrations is intended to maximize detection by creating an extensive library of affinity reagents, which rely on a preserved form of the target protein and might miss genetic effects specific to a precise isoform of the protein. Based on our previous experiments, we collected the media after 24 hours of culture^31,80^. It should be noted that the specific time of collection is crucial as SMC phenotypes undergo changes over time. The production of collagen has been linked to the synthetic behavior of SMCs, as evidenced by various studies^81–83^. It’s worth noting that our studies were conducted *in vitro* and may not accurately reflect the complex interactions that take place *in vivo*. It’s important to recognize that SMCs can undergo phenotypic changes during the onset and progression of vascular diseases, which may not be captured in our *in vitro* system. Lastly, the impact of LTBP1 silencing on the relationship between SMC differentiation and collagen synthesis needs to be elucidated *in vivo*.

In summary, our results contribute to the identification of secreted SMC proteins that may play an important role in vascular diseases. Using these proteome data, we generated the first multi-scale molecular QTL map transcript and protein abundance in SMCs and helped to resolve genetic association signals by identifying likely effector genes.

## Data Availability

All data produced in the present work are contained in the manuscript.

RNAseq data are available at GEO with the accession numbers GSE193817. eQTL results can be accessed at https://virginia.box.com/s/t5e1tzlaqsf85z13o4ie2f9t1i0zfypd and User-friendly website at http://civeleklab.cphg.virginia.edu to query the dataset published in this paper.

## Acknowledgments

This work is dedicated to the late Professor Jeanette Erdmann, a warm-hearted mentor and outstanding researcher whose unique dedication and professionalism greatly shaped an entire field of research. Her legacy will forever be remembered and honored.

The authors thank Sandra Wrobel for her technical support. We also thank the members of the Erdmann, Owens, Kaikkonen and Civelek laboratories for their feedback and discussions.

