## Supplementary Figures for "Secreted protein profiling of human aortic smooth muscle cells identifies vascular disease associations"

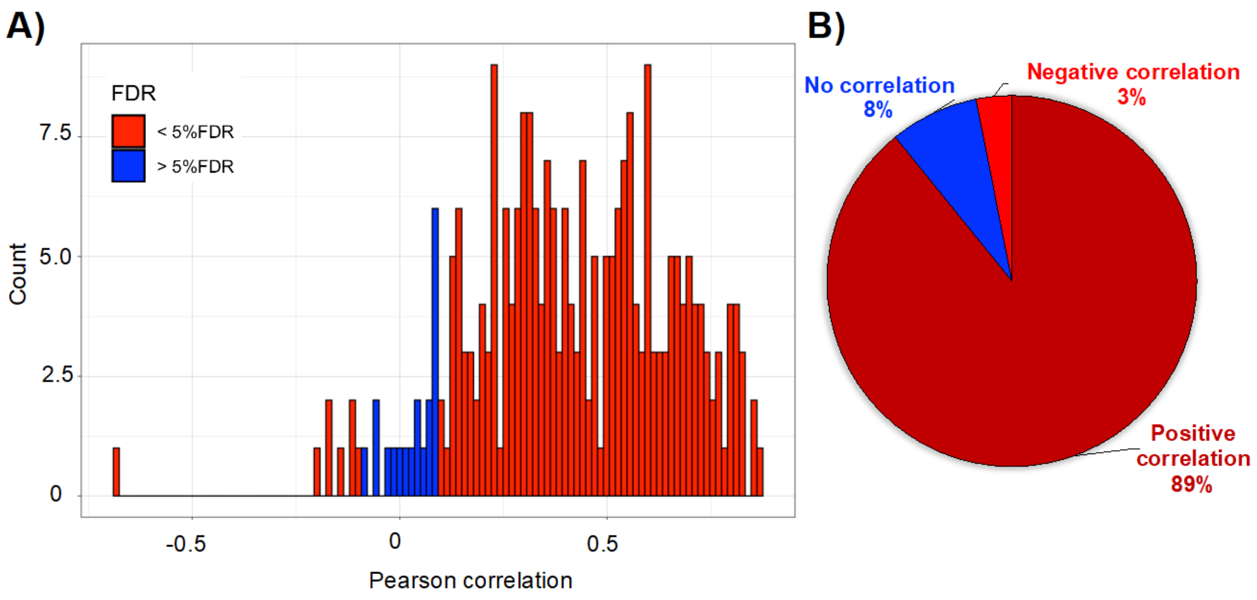


**Supplementary Figure 1: Pearson correlations between the SMC ECM proteins and their transcripts. A)** Histogram of the Person correlations for the 258 (out of 270) ECM protein-transcript pairs. **B)** 89% of the pairwise correlations were positive, 3% of the correlations were negative, and 8% were not significant.

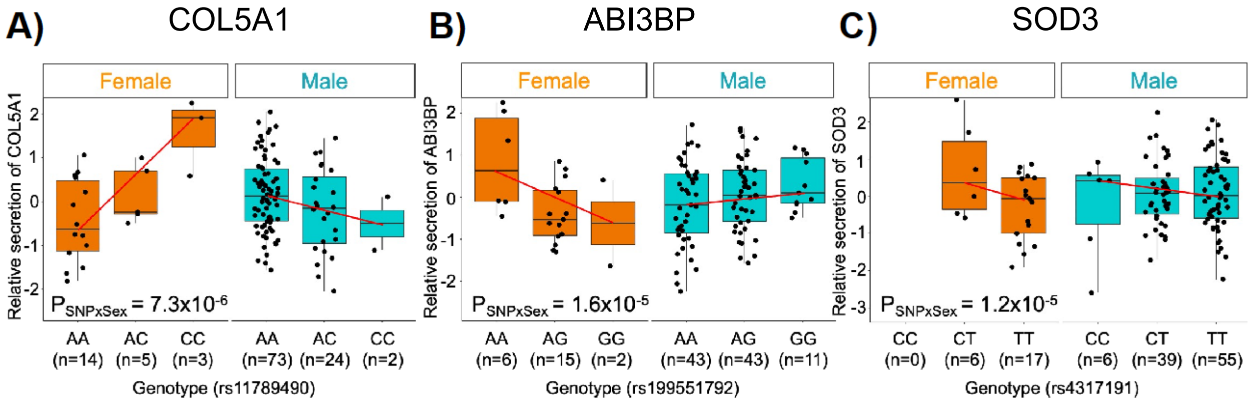


**Supplementary Figure 2: Identification of three sex-biased pQTLs in aortic smooth muscle cells. A-C**) Genotype-protein abundance plots for sex-biased pQTLs for three secreted proteins: **A)** COL5A1, **B)** ABI3BP and **C)** SOD3. The lead variants of the sex-biased pQTLs are shown.

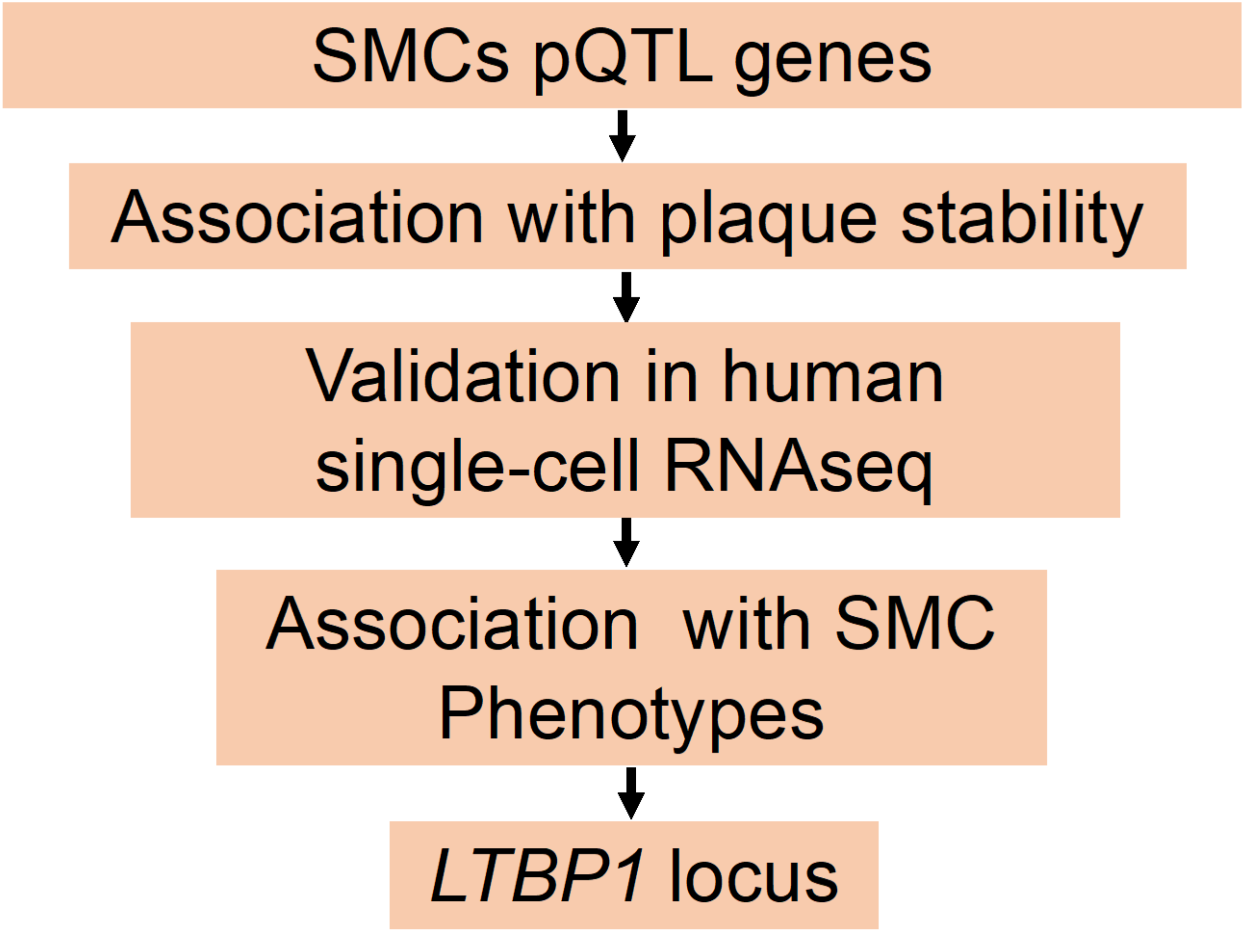


**Supplementary Figure 3: Prioritization criteria of SMC pQTLs for functional studies.**

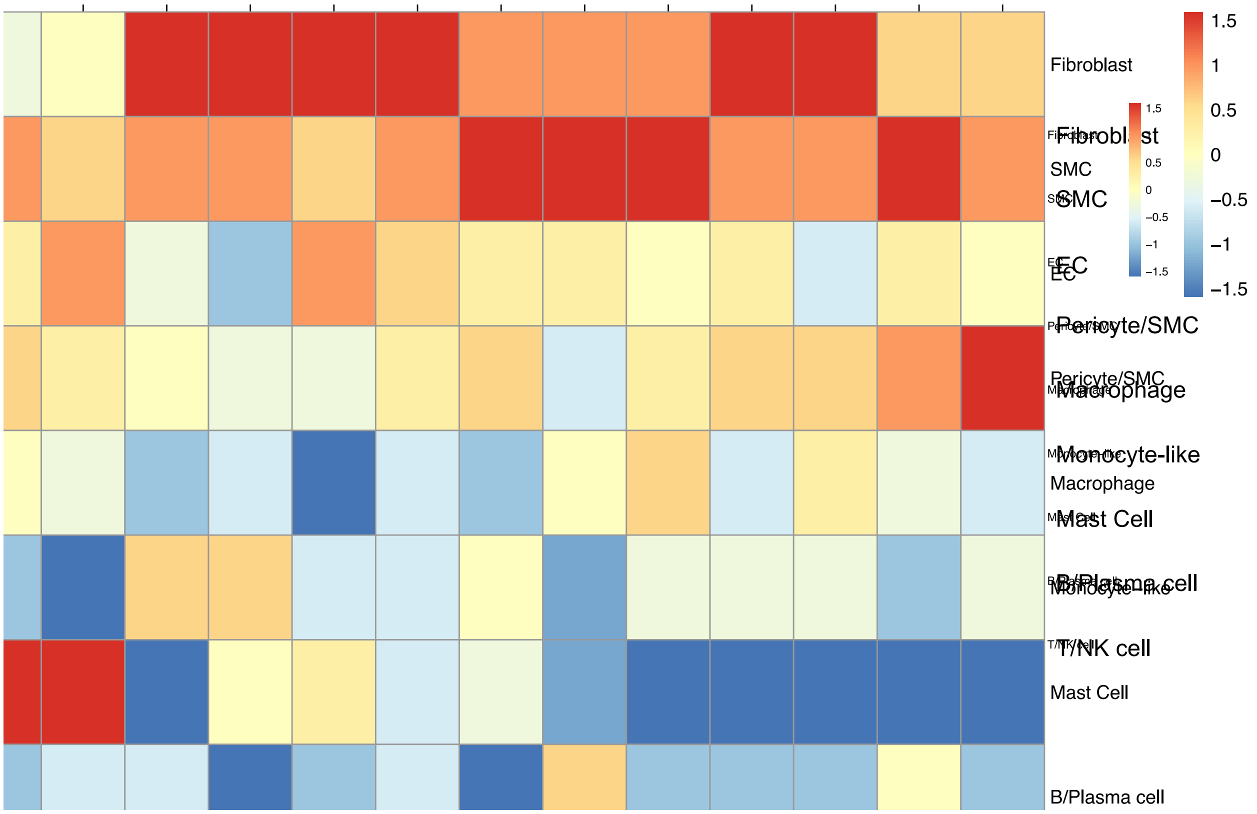


**Supplementary Figure 4: Expression of genes encoding proteins with *cis*-pQTLs in single cells isolated from human coronary atherosclerotic plaques**. The scRNAseq data has been published previously[29]. We were able to assess the expression of 19 of the 20 SMC pQTL genes. 11 of the 19 genes had higher expression in SMCs, pericytes, and fibroblasts compared to endothelial cells, monocytes, macrophages, and other immune cells. The color key of the amount of expression ranging from −1.5 (blue) to +1.5 (red) is shown on the right side. Red color indicates higher expression, while blue color indicates lower expression.

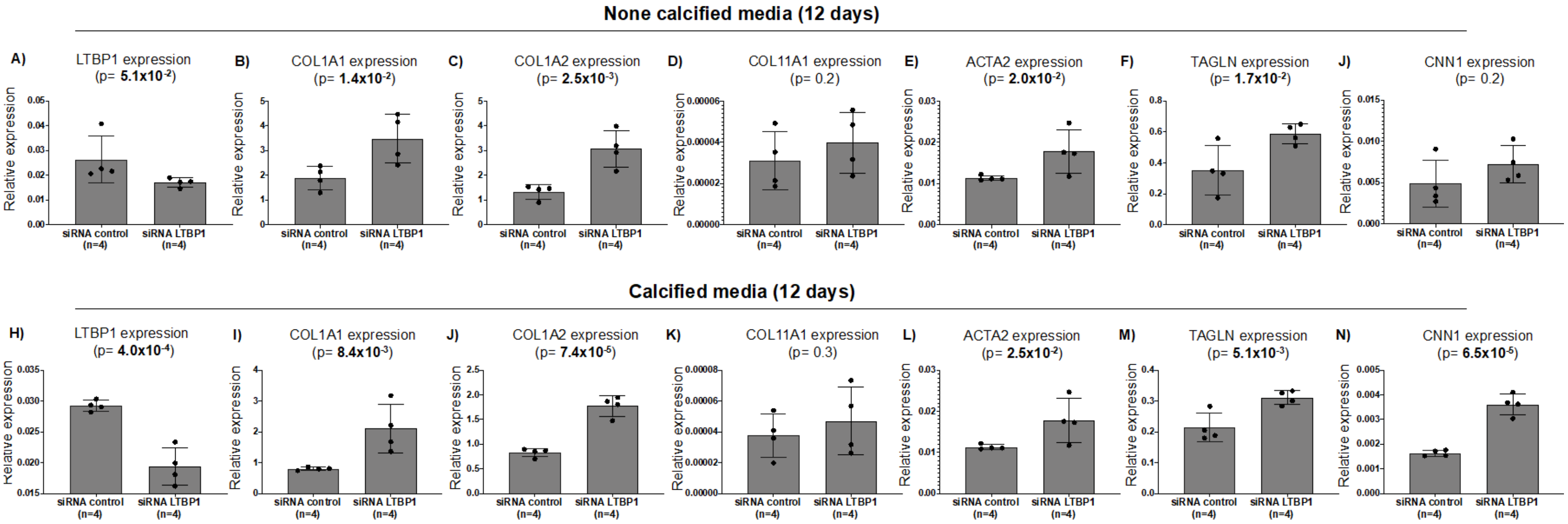


**Supplementary Figure 5: Modulation of *LTBP1* expression in SMCs.** After 12 days of transfection, **A and H)** downregulation of *LTBP1* in SMCs results in a significant increase in the expression of ECM genes both in **B-D)** non-calcified and **I-K)** calcified media. Additionally, SMC-specific marker genes exhibit an increase in expression in **E-J)** non-calcified media and a more pronounced increase in expression in **L-N)** calcified media. n represents the technical repeats from the same donor.

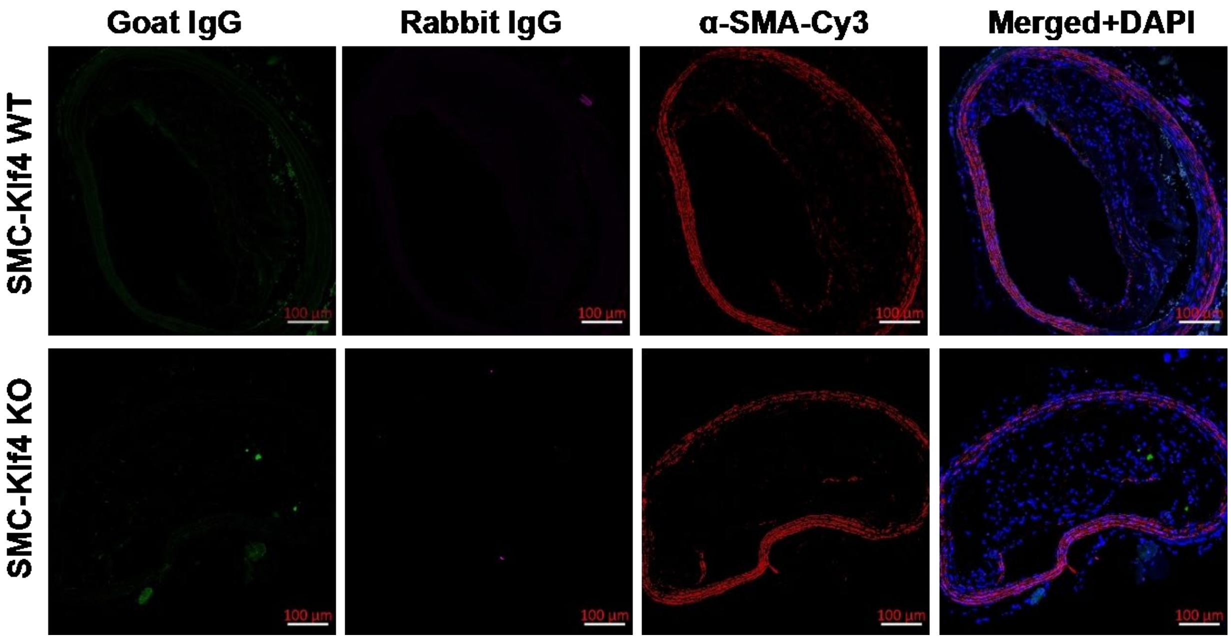


**Supplementary Figure 6: Immunofluorescence images of mouse brachiocephalic artery with isotype control IgG.** Representative immunofluorescence images of brachiocephalic artery lesions from SMC-Klf4 ApoE-/- KO and SMC-Klf4 WT ApoE-/- mice stained for IgG (LTBP1), ACTA2 and IgG (eYFP) after 26 weeks of hypercholesterolemic Western diet.

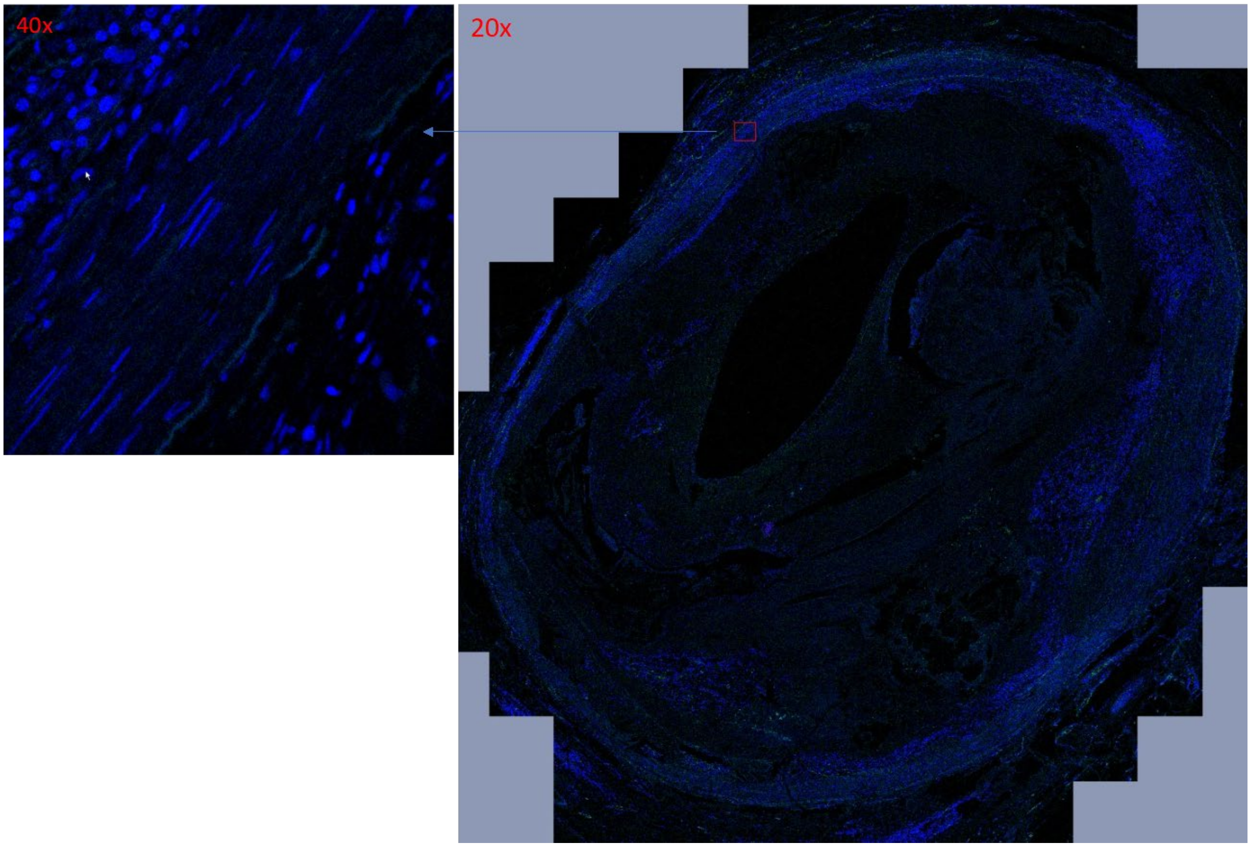


**Supplementary Figure 7**: Immunohistochemistry staining of isotype control IgG. Human fibrous cap atheroma stained for IgG (LTBP1).
